# Covid-19 transmission dynamics during the unlock phase and significance of testing

**DOI:** 10.1101/2020.08.18.20176354

**Authors:** Abhijit Paul, Samrat Chatterjee, Nandadulal Bairagi

## Abstract

The pandemic disease Covid-19 caused by SARS-COV-2, which emerged from Wuhan, China, has established itself as the most devastating disease in the history of infectious disease, affecting 216 countries/territories across the world. Different countries have developed and adopted various policies to contain this epidemic and the most common were the social distancing and lockdown. Though some countries have come out of this pandemic, the infection is still increasing and remains very serious in the rest of the world. Even when the disease is not under control, many countries have withdrawn the lockdown and going through the phase-wise unlocking process, causing a further increment in the infection rate. In such a scenario, the role of the undetected class of infected individuals has become very crucial. The present study is an attempt to understand and estimate the possible epidemic burden during the unlock phase in the presence of an undetected class. We proposed a modified SEIR model and dissected the epidemiological status of different countries with the available data. With the initial establishment of the model with the epidemic data of four countries, which have already attained the epidemic peak, the study focused more on countries like India and the USA, where the epidemic curve is still growing, but the unlock process has started. As a straightforward result, we noticed a significant increase in the undetected and detected infected cases under the ongoing unlock phase. Under such conditions, our recalibration exercise showed that an increase in the testing could revert the existing growth rate of the infected cases to the lower growth rate of the lockdown period. Our present study emphasizes on the implementation of 3T principles, trace, test, and treat, to contain the epidemic. The significance of large scale testing in controlling the epidemic is true for both India and the USA though they have different socio-economic conditions. The use of repurposing drugs may further decrease the infected cases and help the disease controlling process. We believe our proposed strategy obtained through a mathematical model will help to make a better policy for the unlock phase.

## 1 Introduction

The coronavirus disease 2019 (Covid-19), which originated from Wuhan, China, and declared as a pandemic by the World Health Organization (WHO) on March 11, 2020 [1], has put several countries under total or partial lockdown. After observing significant days of lockdown, many countries either have withdrawn or are planning to withdraw the lockdown in phases, even 39 though the epidemic situation is not conducive and the aftermath of withdrawal is mostly unknown. Following the advice of WHO [2], health administrators are paying more attention to large scale testing to trace the symptomatic and asymptomatic covid positive cases [5, 6, 7]. Many countries have followed this instruction and intensified the covid testing, which has helped to trace and isolate the infectives more quickly so that the spreading chain can be discontinued [3]. Country-wise data on the testing till 22nd May showed that Spain and Italy crossed 50,000 tests per million population, while the USA, Germany and Switzerland crossed more than 40,000 tests per million population [3]. However, most of the African, South American and Asian countries including India have a testing rate of less than 15,000 per million or less, which is significantly low and is a hurdle in the prevention and control measures [4]. This is because of the shortage of test kits, face masks and PPE (personal protection equipment) across the world and high cost of testing kits, causing an increase in the undetected cases of Covid-19 due to non-identification [7, 8, 9]. Such undetected classes of infected individuals might play an important role in the disease spreading and posing a challenge in the containment policies of the government [10, 11, 12]. This class will certainly be more influential during the unlock period because of their unrestricted movement. The present study is an attempt to understand and estimate the possible epidemiological burden of a country during the unlock period in the presence of undetected class and discuss some policy to combat the infection spreading during the unlock period.

The trend of an epidemic can be better understood, even in the case of an unusually fast outbreak like Covid-19, through mathematical models. Several such mathematical models [13, 14, 15, 16, 17, 18] have been proposed and analyzed for Covid-19 pandemic since its outbreak. These models used available data to predict the expected cases in the near future and the number of deaths. The prediction about the epidemic peak, cases during the peak periods, and the information about its duration help policy-makers to make necessary plans to fight against the epidemic. Some models were made for multiple countries [10, 19] and some were focused on specific countries like China [11], Cameroon [12], Italy [20, 21] and India [22, 23, 24, 25, 26]. These studies considered different models on early Covid-19 transmission dynamics and predicted the epidemic burden as well as the future of the diseases in the presence or absence of lockdown. The current study differs from the mentioned studies and considers mainly the epidemic status during the unlock phase. As mentioned earlier, the undetected positive individuals may change the epidemiological burden and are crucial from the management viewpoint. We are, therefore, kin to know the effect of such class in the unlock phase and find a way out to reduce the number of infectives during this period. In this process, we first aimed to understand the effect of lockdown in controlling the pandemic in countries that attained the epidemic peak and where the number of daily cases is going down. We then studied the Covid 19 disease dynamics of countries like India and the USA, where the epidemic curve is still growing even after implementing a longer lockdown, to identify the possible reasons for not flattening the curve. The undetected infectives are possibly one of the main reasons for making the policymakers upset as the outcome from the intervention measures was far from the expectation. The second aim of the current study is, therefore, to dissect the possible influence of the undetected infective class on the disease dynamics during the unlock period and make some policy hypotheses to combat the infection spreading during this period.

The paper is divided into several sections with model construction in the immediate next Section 2. Section 3 deals with the country-wise model analysis. In Section 4, we studied the significance of undetected infected individuals in the spreading of Covid-19. We looked for possible policy hypotheses through parameter recalibration in Section 5. The article ends with a discussion in Section 6.

## 2 The 86 mathematical model

Our proposed model is based on the classical SEIR (susceptible, exposed, infectious, recovered) epidemic model. However, an extension in this basic model was done to incorporate other compartments in the population based on the characteristics of Covid-19 infection. Depending on the epidemiological status of an individual, we classified the population into susceptible (*S*), exposed (*E*), detected infectious (*I_d_*), undetected infectious (*I_u_*), recovered from the detected class (*R_d_*) and recovered from the undetected class (*R_u_*). Besides, to give accountability on the virulence of the disease, deaths from the detected class (*D_d_*) and deaths from the undetected class (*D_u_*) have also been considered.

Each individual is assumed to be susceptible to SARS-COV-2 because it is a novel virus and there is no immunity against it. It is reported that a large portion of Covid positive individuals do not show any symptoms or show mild symptoms [27, 28], and a portion of such asymptomatic covid positive individuals remains undetected [29, 30]. It is to be mentioned that an asymptomatic individual may be a member of *I_d_* class if Covid-19 infection is confirmed by rapid antigen or PCR tests, but no member of *I_u_* class has gone through test. Susceptible individuals (*S*) get infection from both the detected (*I_d_*) and undetected (*I_u_*) infectious individuals. Detected Covid positive individuals are assumed to spread infection at a lower rate as they are identified, and this fact is incorporated in the incidence rate by a scaling parameter *α* (0 *< α <* 1). The disease incidence rate or the rate of new infection is assumed to follow nonlinear incidence, as it better fits the epidemiological data compared to bilinear incidence [31], with *β* as the disease transmission coefficient. All newly infected individuals join the exposed class *E*, who carry the virus but do not spread infection, and individuals leave this class at a rate *ξ* after spending an average time^1^_*ξ*_ units (day). Note that the median period in staying *E* class is 3.69 day [32], giving the value of *ξ* in between 0 and 1. A fraction γ (0 < γ < 1) of them join *I_d_* class and the remaining 110 fraction (1 *− γ*) becomes the member of *I_u_* class. An individual of undetected class may join the detected class if tested positive later on, and such transformation from *I_u_* class to *I_d_* class may occur at a rate *ω*. Transformation from *I_d_* class to *R_d_* class through recovery occurs at a rate *δ_d_* and a similar transformation rate from *I_u_* class to *R_u_* class is denoted by *δ_u_*. The disease related death rates in the *I_d_* and *I_u_* classes are denoted, respectively, by *µ_d_* and *µ_u_*, and these individuals join the respective death classes *D_d_* and *D_u_*. The parameters *δ_d_, δ_u_, µ_d_, µ_u_* and *ω* all lie in between 0 and 1. We assume that the total population (*N*) remains constant throughout the study period, a valid assumption if the epidemic period is not too long. The interaction among different classes with these assumptions may be represented by the following system of differential equations:

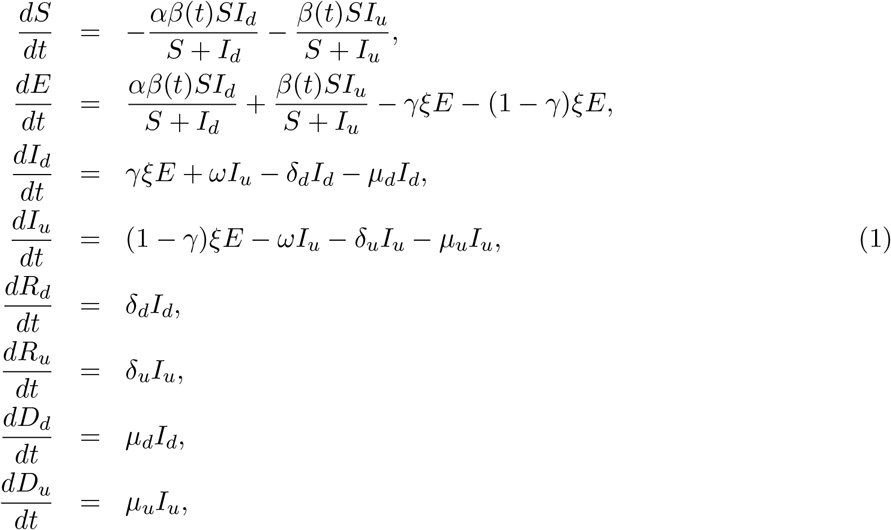

where

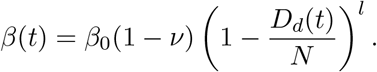

Here *β*_0_(1 *− ν*) is the disease transmission efficiency in the presence of lockdown with *β*_0_ as the baseline disease transmission rate. In fact, the parameter *ν* encapsulates the effect of lockdown. Thus, *ν* takes the value 0 if there is no lockdown, otherwise 0 *< ν <* 1. Public perception of getting an infection may act negatively on disease transmission. This perception grows with increasing corona-virus-related death [33, 34]. The last expression accounts for such perception and affects negatively on the disease transmission due to human behavioral responses to deaths, where *l* measures the strength of this response.

### Basic reproduction number

The basic reproduction number determines whether an infection will spread in the community or not. It is the average number of secondary cases produced by an index case. If this number is greater than 1 then an epidemic can grow, otherwise it dies out [35]. Close to the disease-free equilibrium (DFE) of model (1), a state in which S = N (*N* being the constant population size) and *E* = *I_d_* = *I_u_* = 0, the Jacobian matrix of the infection subsystem *{E, I_d_, I_u_}* is *J*_0_ = *F − V*, where *F* is the transmission matrix and V is the transition matrix. The basic reproduction number (*R*_0_) is the spectral radius of the next-generation matrix (*NGM*) [36], where *NGM* = *F V*^−1^.

Here

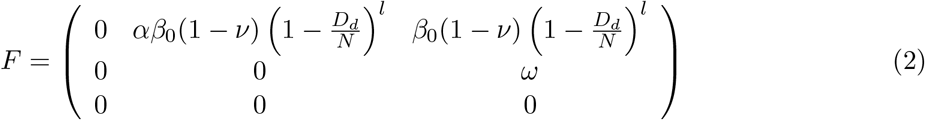

and

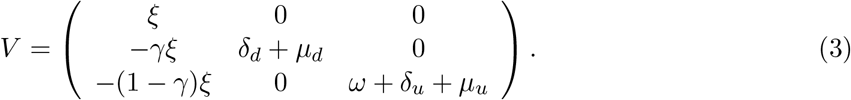

The next generation matrix (*NGM*) then can be found as

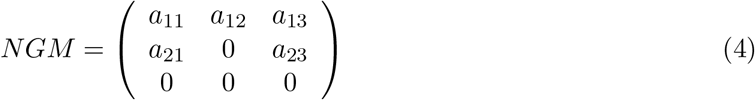

with

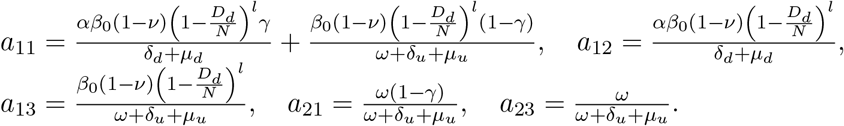

The Characteristic polynomial of this *NGM* is

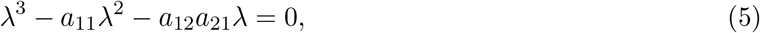

having roots 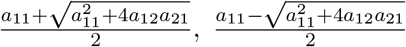 and 0. Noting that *a*_11_, *a*_12_ and *a*_21_ are always positive, as because of 0 *< ν <* 1, 0 *< γ <* 1 and *D_d_ < N*, the only positive root of (5) is 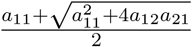. Therefore, following [36], the basic reproduction number of our system is *R*_0_ = 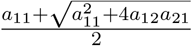.

## 3 Simulation results

The country-wise temporal data of Covid-19 disease was taken from a web-based interactive dash-board maintained by the Centre for Systems Science and Engineering (CSSE) at Johns Hopkins University [37, 38]. The rationale to develop this type of dashboard is to visualize and track reported cases of Covid-19 in real-time and make these data freely available for knowledge development. We have used the country-wise Covid-19 outbreak data of confirmed, recovered, and death cases till July 27, 2020. We first considered the data from Italy, Spain, Germany, and Switzerland, where the epidemic curve has been flattened due to the implementation of strict containment policies. Subsequently, we studied countries like India and the US, where unlocking has started without achieving the epidemic peak.

### 3.1 Analysis for Italy, Spain, Germany, and Switzerland

#### Parameter estimation

It was difficult to find a single parameter set that fits the data of the entire epidemic period of a country because lockdown affected the disease spreading and consequently some model parameters significantly changed their values in the pre-lockdown, lockdown, and post-lockdown periods. We, therefore, partitioned the study period into three-time segments, namely BLS (before lockdown stage), ELS (early lockdown stage), and LLS (late lockdown stage), using two critical time-points (see Supplementary Information and Supplementary Methods S1 for justification and technique). Only five parameters were selected through global sensitivity analysis (see Supplementary Methods S4 and Supplementary Figure S2) which would be varied in the three-time segments keeping other parameters fixed. The optimal parameter sets (see Table 1) were then generated for three-time segments using ”lsqcurvefit” function from the Optimization Toolbox of MATLAB and by mean normalized Euclidean distance (MNED) technique (see Supplementary Methods S2 and S3). With these optimal parameters, the model (1) was simulated for each country and the results were compared with the real data (see Figure 2).

**Table 1:** Optimal parameters for BLS, ELS, and LLS time segments, separated by the two critical time points, for the countries Italy, Spain, Germany, and Switzerland. Parameters *l, γ, ξ, ω, δ_u_, µ_u_* remained fixed for each time segment, while the other five parameters differed. The first critical time points are March 10, 14, 23, and 17, respectively, the lockdown starting dates for Italy, Spain, Germany, and Switzerland. The second critical time points for these countries are 37 days, 23 days, 14 days, and 15 days after the first critical point. The population of a country was considered as the initial value of the susceptible population of that country.

|  | Country |  |  |  |
| --- | --- | --- | --- | --- |
| Parameters/<br>initial values | Italy | Spain | Germany | Switzerland |
| $l$ | 8 | 8 | 4 | 5.6 |
| $\gamma$ | 0.281 | 0.576 | 0.294 | 0.204 |
| $\xi$ | 0.167 | 0.66 | 0.627 | 0.66 |
| $\omega$ | 0.25 | 0.305 | 0.277 | 0.133 |
| $\delta_u$ | 0.016 | 0.066 | 0.02 | 0.037 |
| $\mu_u$ | 0.0015 | 0.006 | 0.001 | 0.0023 |
| **Before lockdown stage (BLS)** |  |  |  |  |
| $\alpha$ | 0.23 | 0.1 | 0.21 | 0.18 |
| $\beta_0$ | 1.48 | 1.42 | 0.63 | 1.3 |
| $\nu$ | 0 | 0 | 0 | 0 |
| $\delta_d$ | 0.036 | 0.013 | 0.0029 | 0.00076 |
| $\mu_d$ | 0.03 | 0.012 | 0.0007 | 0.0036 |
| **Early lockdown stage (ELS)** |  |  |  |  |
| $\alpha$ | 0.085 | 0.085 | 0.06 | 0.164 |
| $\beta_0$ | 0.49 | 1.2 | 0.55 | 0.33 |
| $\nu$ | 0.47 | 0.41 | 0.42 | 0.49 |
| $\delta_d$ | 0.013 | 0.052 | 0.043 | 0.016 |
| $\mu_d$ | 0.0096 | 0.016 | 0.002 | 0.0043 |
| **Later lockdown stage (LLS)** |  |  |  |  |
| $\alpha$ | 0.05 | 0.34 | 0.06 | 0.067 |
| $\beta_0$ | 0.4 | 0.32 | 0.45 | 0.33 |
| $\nu$ | 0.82 | 0.86 | 0.565 | 0.754 |
| $\delta_d$ | 0.032 | 0.024 | 0.074 | 0.076 |
| $\mu_d$ | 0.0025 | 0.0028 | 0.0038 | 0.0038 |
| **Initial values of the variables** |  |  |  |  |
| $S_0$ | 6,03,17,546 [42] | 4,67,33,038 [43] | 8,30,42,200 [44] | 85,70,146 [45] |
| $E_0$ | 0 | 0 | 0 | 0 |
| $I_{d0}$ | 2 | 1 | 1 | 1 |
| $I_{u0}$ | 0 | 0 | 0 | 0 |
| $R_{d0}$ | 0 | 0 | 0 | 0 |
| $R_{u0}$ | 0 | 0 | 0 | 0 |
| $D_{d0}$ | 0 | 0 | 0 | 0 |
| $D_{u0}$ | 0 | 0 | 0 | 0 |

**Figure 1:**
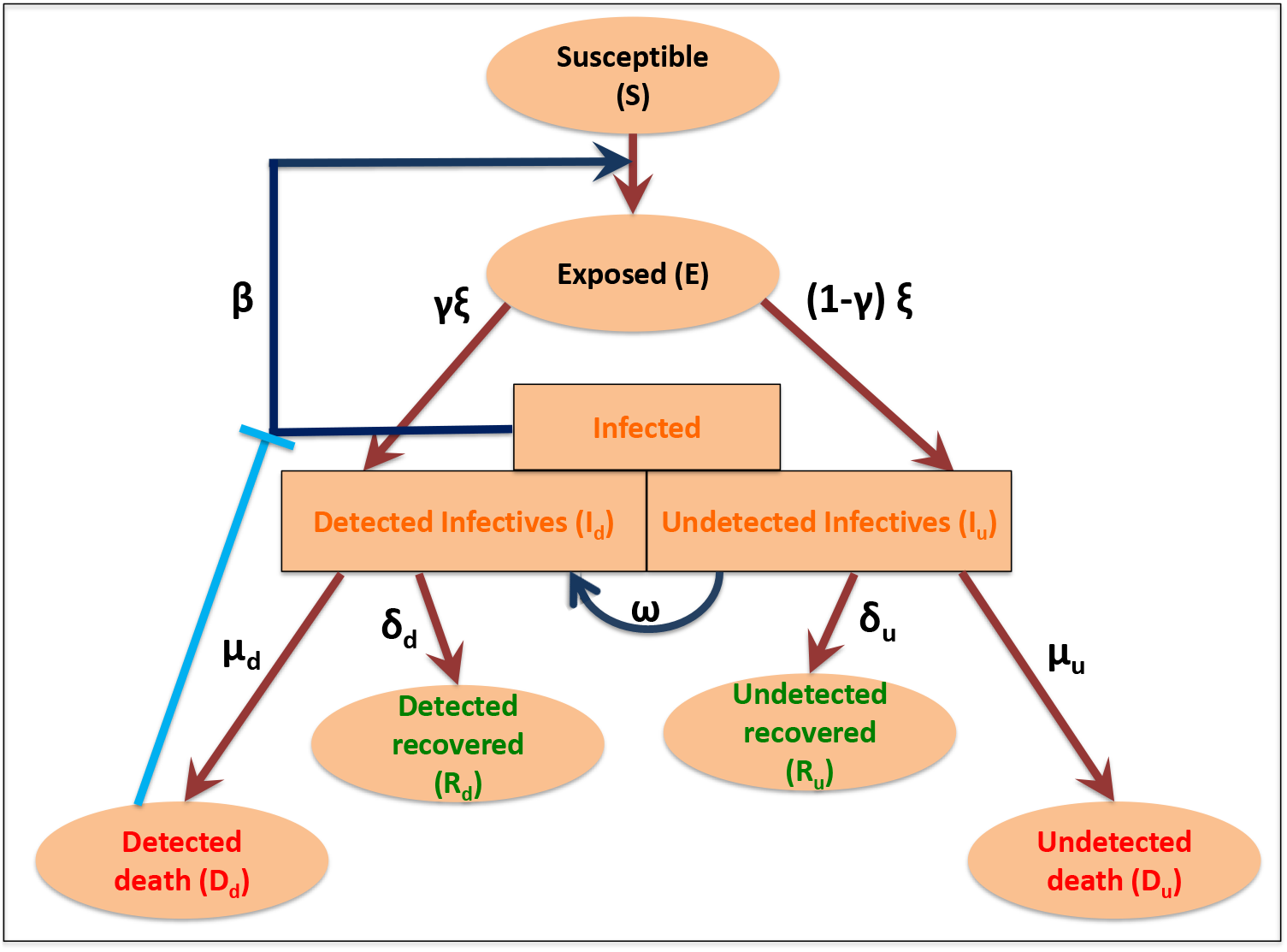
Schematic diagram of our proposed Covid-19 epidemic model.

**Figure 2:**
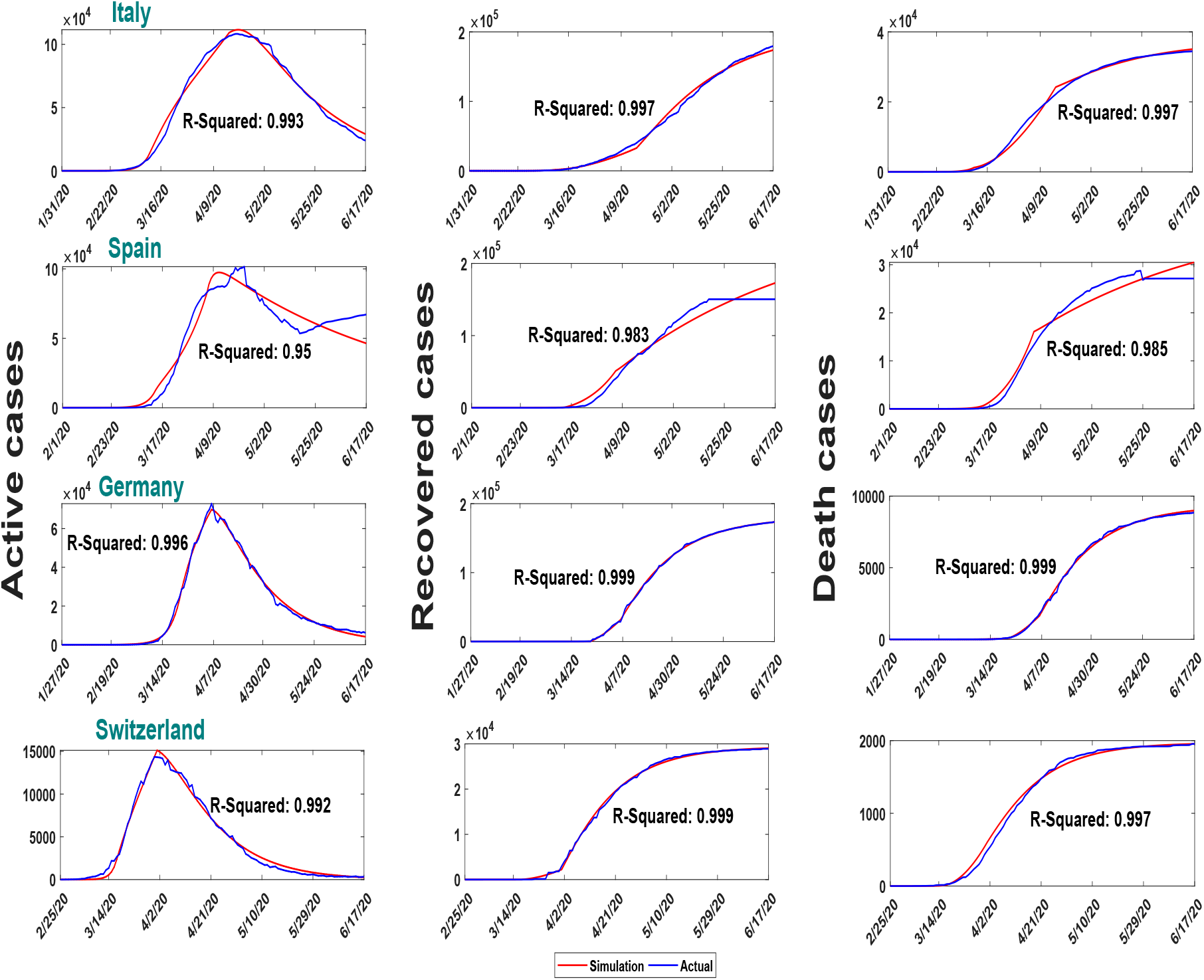
Country-wise comparison of real data (blue colour) of active, recovered and death cases with their corresponding simulated data (red colour) for the period January 22 to June 27, 2020. Parameters are as in Table 1. Data fitting was relatively poor in the case of Spain because they changed the methodology on April 19 and May 25. In these day daily cases were shown to be negative, and the cumulative cases became lower than its immediate previous day.

#### 3.1.1 Lockdown effect on the disease transmission rate (*β*(*t*)) and basic reproduction number (*R*_0_)

The objective of lockdown is to reduce the transmission rate of the disease, represented by *β*(*t*), so that infection does not spread in the community. On the other hand, the health of the community is measured by the transmission potentiality measuring parameter, *R*_0_. We, therefore, observed the effect of lockdown on the parameter *β*(*t*) and the index parameter *R*_0_. We plotted (Figure 3) the variation in the values of *β*(*t*) and *R*_0_ with respect to time. It shows that both *β* and *R*_0_ were significantly high for all four countries in the BLS period and then it gradually decreased below 1 in the LLS, implying that the disease was controlled in this period.

**Figure 3:**
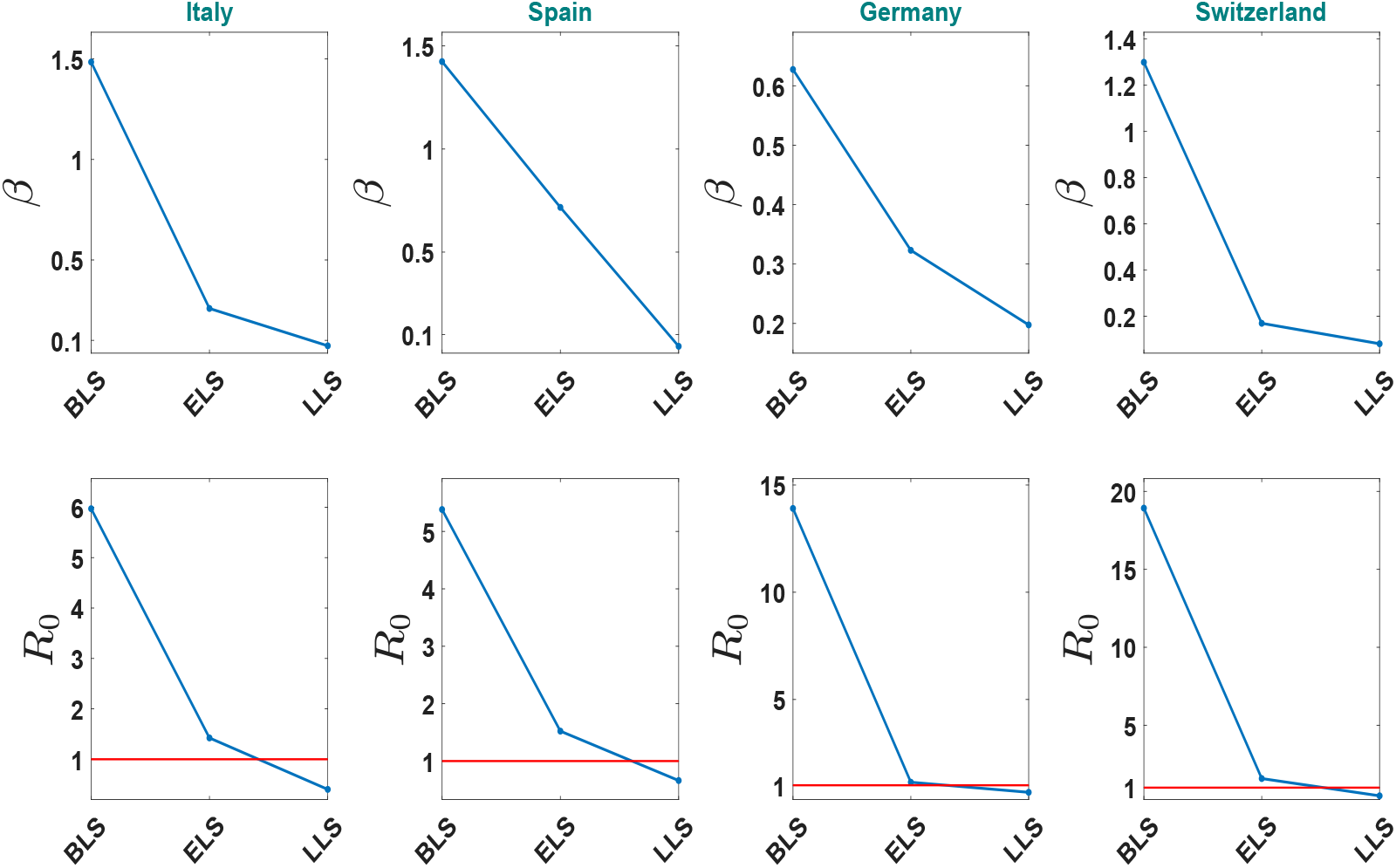
Variation in the disease transmission rate, *β*(*t*), and the basic reproduction number, *R*_0_, of each country with respect to time. The critical line *R*_0_ = 1 is represented by red color. Epidemic will be controlled if *R*_0_ goes below this line.

### 3.2 Analysis for India and US

#### Parameter estimation

The lockdown starting dates for India and the US are 25th March and 20th March, respectively, [40] and these dates were used as their first critical time-point. Following the earlier protocol, we generated the optimal parameter set (Table 2) for India and USA (see Supplementary Information). With these parameters, the model (1) was simulated and the results were compared with the real data of India and the USA (see Figure 4). We here used an additional time segment ULS (unlock stage) because we wanted to compare between two estimated values of infectives under uncontrolled and partially controlled situations during this unlock period.

**Table 2:** Optimal parameters for BLS, ELS, LLS and ULS time segments, separated by the three critical time points, for India and USA. Parameters *l, γ, ξ, ω, δ_u_, µ_u_* remained fixed for each time segment, while the other five parameters differed. The first critical time points are March 25 for India and March 20 for the USA, which are the lockdown starting dates of the two countries. The second critical time points for these countries are 33 days and 45 days, after the first critical point. The third critical point was the unlocking day and it is July 1 for India [46, 47] and July 14 for USA (New York) [40]. The initial value of the susceptible population was considered as the population of the respective country.

| Parameters/ initial values | Country |  |
| --- | --- | --- |
|  | India | US |
| $l$ | 10.5 | 12 |
| $\gamma$ | 0.34 | 0.34 |
| $\xi$ | 0.26 | 0.34 |
| $\omega$ | 0.33 | 0.2 |
| $\delta_u$ | 0.04 | 0.033 |
| $\mu_u$ | 0.0012 | 0.002 |
| **Before lockdown stage (BLS)** |  |  |
| $\alpha$ | 0.07 | 0.12 |
| $\beta_0$ | 1.07 | 0.92 |
| $\nu$ | 0 | 0 |
| $\delta_d$ | 0.005 | 0.0028 |
| $\mu_d$ | 0.0028 | 0.0067 |
| **Early lockdown stage (ELS)** |  |  |
| $\alpha$ | 0.18 | 0.095 |
| $\beta_0$ | 0.659 | 0.498 |
| $\nu$ | 0.41 | 0.41 |
| $\delta_d$ | 0.026 | 0.0071 |
| $\mu_d$ | 0.0033 | 0.0044 |
| **Later lockdown stage (LLS)** |  |  |
| $\alpha$ | 0.228 | 0.155 |
| $\beta_0$ | 0.62 | 0.365 |
| $\nu$ | 0.529 | 0.724 |
| $\delta_d$ | 0.048 | 0.0087 |
| $\mu_d$ | 0.0023 | 0.0013 |
| **Unlock stage (ULS)** |  |  |
| $\beta_{0_1}$ | 0.62 | 0.365 |
| $\nu_1$ | 0.529 | 0.724 |
| $\beta_{0_2}$ | 0.659 | 0.498 |
| $\nu_2$ | 0.41 | 0.41 |
| $\gamma$ | 0.37 | 0.33 |
| $\omega$ | 0.5 | 0.3 |
| $\delta_d$ | 0.059 | 0.008 |
| $\delta_u$ | 0.06 | 0.05 |
| $\mu_d$ | 0.002 | 0.00086 |
| $\mu_u$ | 0.0018 | 0.003 |
| **Initial values of the variables** |  |  |
| $S_0$ | 135,26,42,280 [48, 49] | 32,92,27,746 [50] |
| $E_0$ | 0 | 0 |
| $I_{d_0}$ | 1 | 1 |
| $I_{u_0}$ | 0 | 0 |
| $R_{d_0}$ | 0 | 0 |
| $R_{u_0}$ | 0 | 0 |
| $D_{d_0}$ | 0 | 0 |
| $D_{u_0}$ | 0 | 0 |

**Figure 4:**
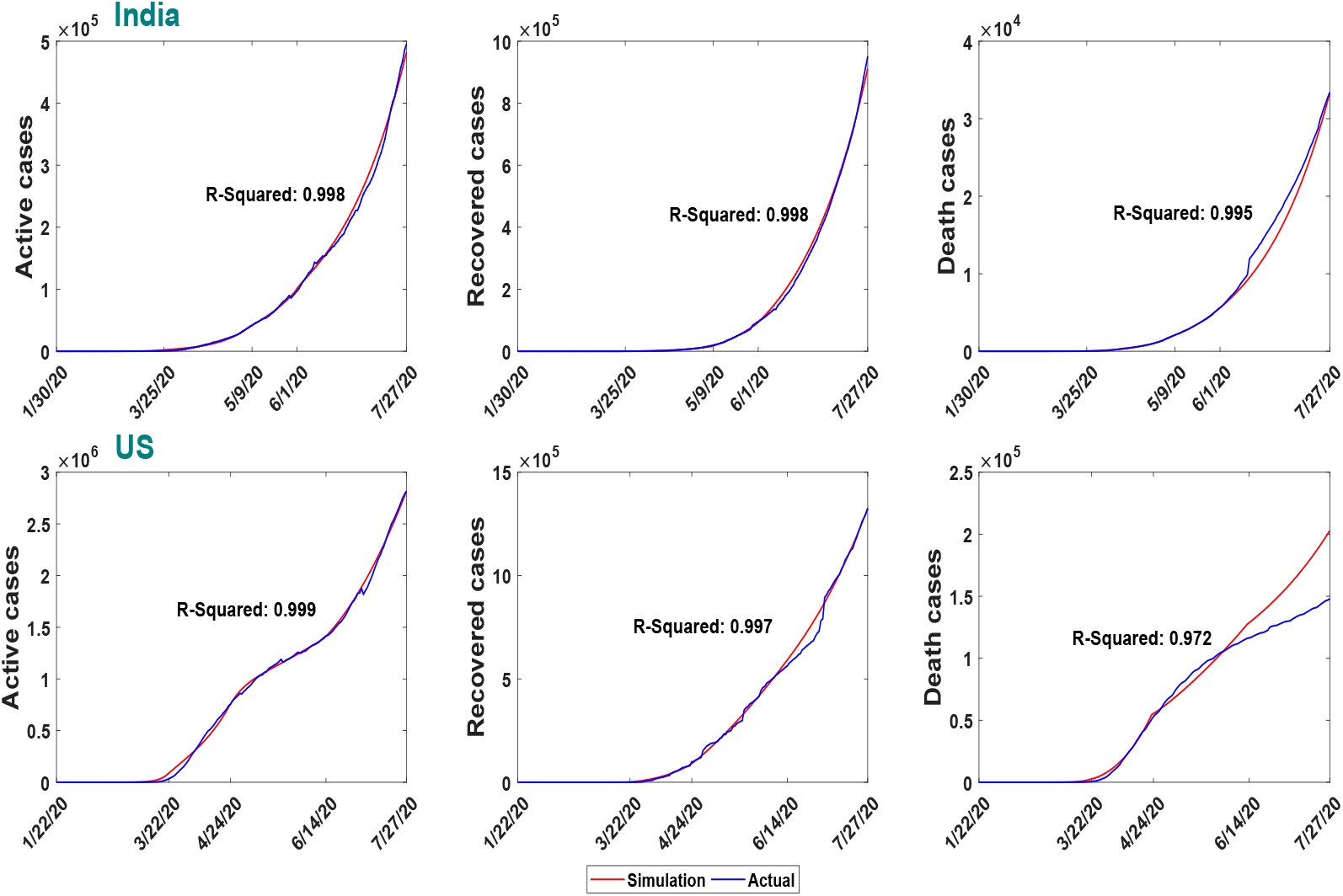
Comparison between the model simulation data and the reported data of India and the USA. The data contains case information for the period January 30 to July 27 for India and January 22 to July 27 for USA. The parameter values for the simulation are given in Table 2. Three critical time points that separate the study period are mentioned in the labels of the figures.

##### 3.2.1 Lockdown effect on the disease transmission rate (*β*(*t*)) and basic reproduction number (*R*_0_) of India and USA

To observe the effect of lockdown on the disease transmission rate (*β*(*t*)) and the basic reproduction number of India and USA, we plotted their variation (see Figure 5) for three-time stages as before. During BLS, these values were very high for both the countries, but it decreased later on and remained in between 1 and 2 for rest of the period, implying that the disease is still not under control. Note that the transmission rate increased in both countries during the unlock period.

**Figure 5:**
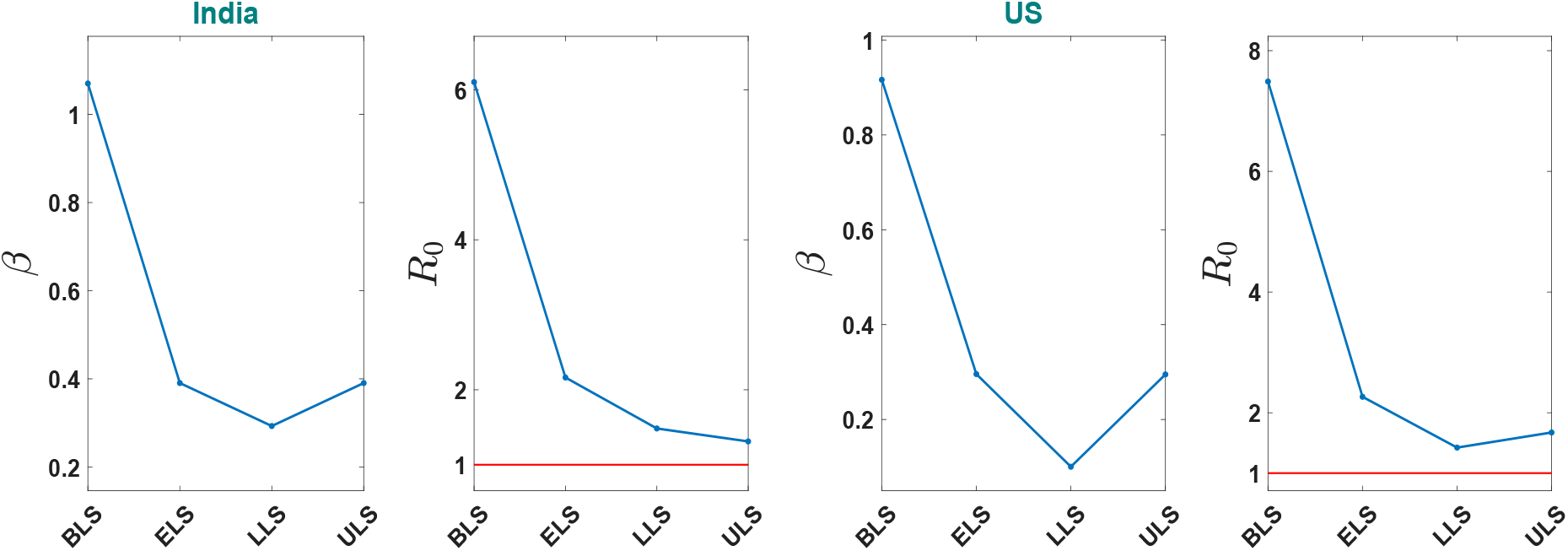
Variation in the disease transmission rate, *β*(*t*), and the basic reproduction number, *R*_0_, of India and USA with respect to the time. The disease is still growing as the basic reproduction number is above the line of control *R*_0_ = 1 (red color line).

## 4 Significance of undetected infected individual class in the spreading of Covid-19

### Estimating country-wise undetected infected individual

It is reported that 81% of Covid-19 infected individuals are mildly symptomatic or asymptomatic [51]. Unfortunately, a significant number of asymptomatic cases remain undetected due to the limitation of testing and unawareness of the population [32]. To capture the effect of the undetected infected population in the spreading of disease, we plotted the time-series curve for the undetected infected class for all six countries in Figure 6. It shows that European countries were able to reduce the undetected cases with the controlling of disease, but the number is still increasing in India and USA. Though there was a negative slope in the undetected infectives curve of USA during April end to May middle, USA couldn’t hold it up, and the number surged again.

**Figure 6:**
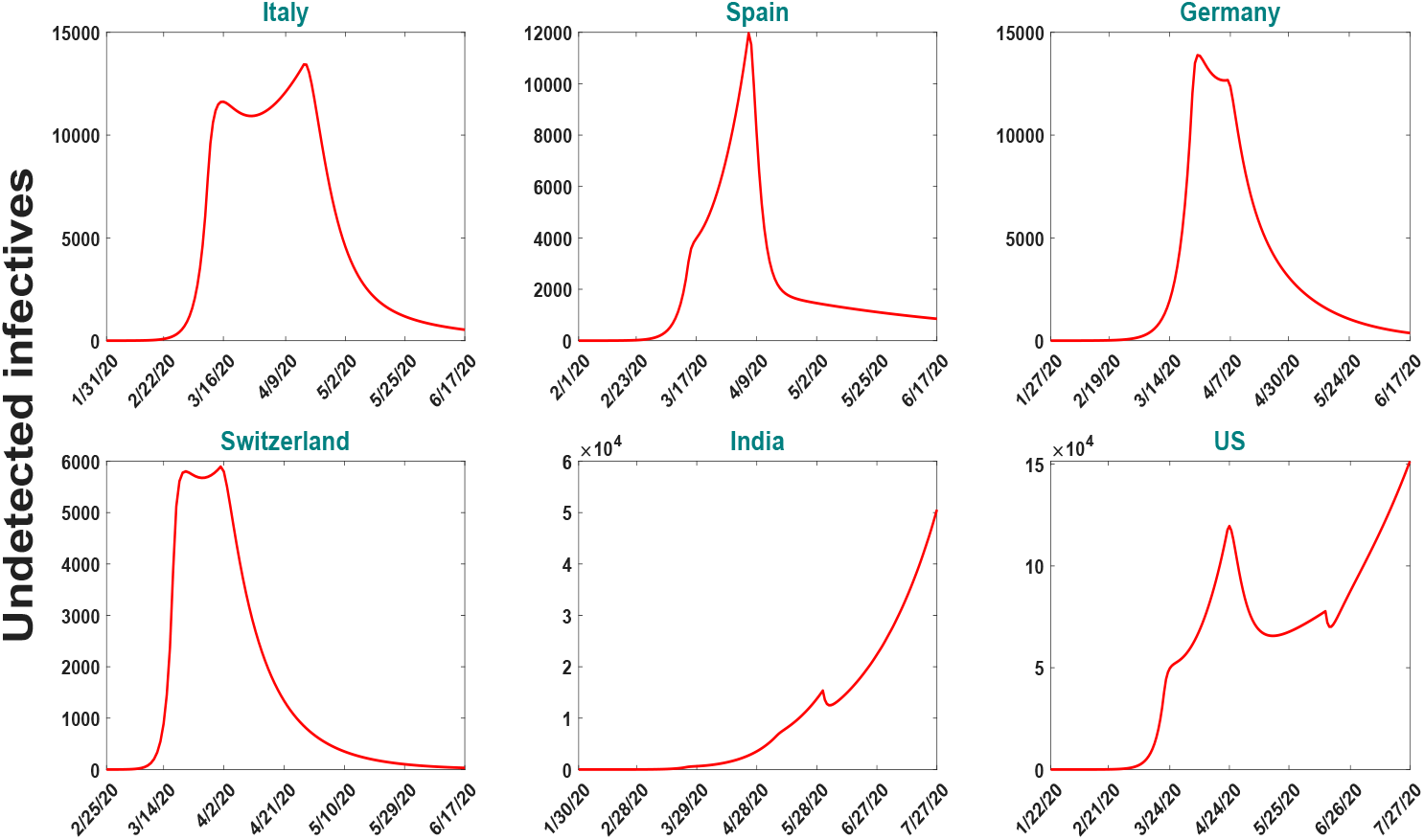
Time evolutions of the undetected class (*I_u_*) for the six nations with parameter values given in Table 1 and Table 2 with their respective time period. The number of individuals in this class has decreased in all countries except India and USA.

### Prediction on the detected and undetected infected class

The aim here is to catch the impact of undetected infected class on the disease spreading during the unlock phase in case of India and the US. As most of the undetected covid positive individuals are asymptomatic or mildly symptomatic, they do everything as healthy individuals do. During the unlock period, there will be no restriction in the movement and consequently, these undetected people would play a pivotal role in spreading the infection. There would be a high chance of community spreading during this period and contact tracing will very difficult, even may be impossible in many cases. So the question is how can we minimize the undetected cases and restrict spreading during this unlock period? As mentioned earlier, some parameter values are expected to change in the unlock period compare to their lockdown values, which would contribute to worsening the epidemic situation in the unlocking stage. So a pertaining question is – what changes we may expect in the detected and undetected classes if the parameter values during the unlock period are changed to the parameter values of the LLS condition? For such comparison study, we plotted the fold change in the levels of *I_d_, R_d_, D_d_, I_u_, R_u_* and *D_d_* after 1 month, 2 months and 3 months considering the current values as their respective base values (see Figure 7). The red color bars in Figure 7 represent the fold change in each class if there is no change in the parameter values of ULS period and the blue color bars represent the fold change with respect to its July 27 value if the parameters take the value of the LLS period instead of ULS. For example, if the parameter values of ULS condition is maintained then there will be a 2.26 times increase in the *I_d_* class in comparison to that of July 27 value. However, if the parameter values are changed to LLS condition from July 28 then the fold change in *I_d_* after one month will be 1.86 times the corresponding value of July 27. Such fold change will be 5 and 11.8 times the value of July 27 in the *I_d_* class after two and three months, respectively, if parameters in the ULS condition are maintained. However, if the parameters are changed to LLS condition then the corresponding fold change will be 3.2 and 6 times the value of July 27. Thus, there would be a huge change in every compartment after three months of unlocking period if the parameter values can be changed to LLS stage. Similar changes are also observed in the case of the USA. In the next subsections, we demonstrate how to change the parameter values of the unlock period to the parameter values of the LLS period.

**Figure 7:**
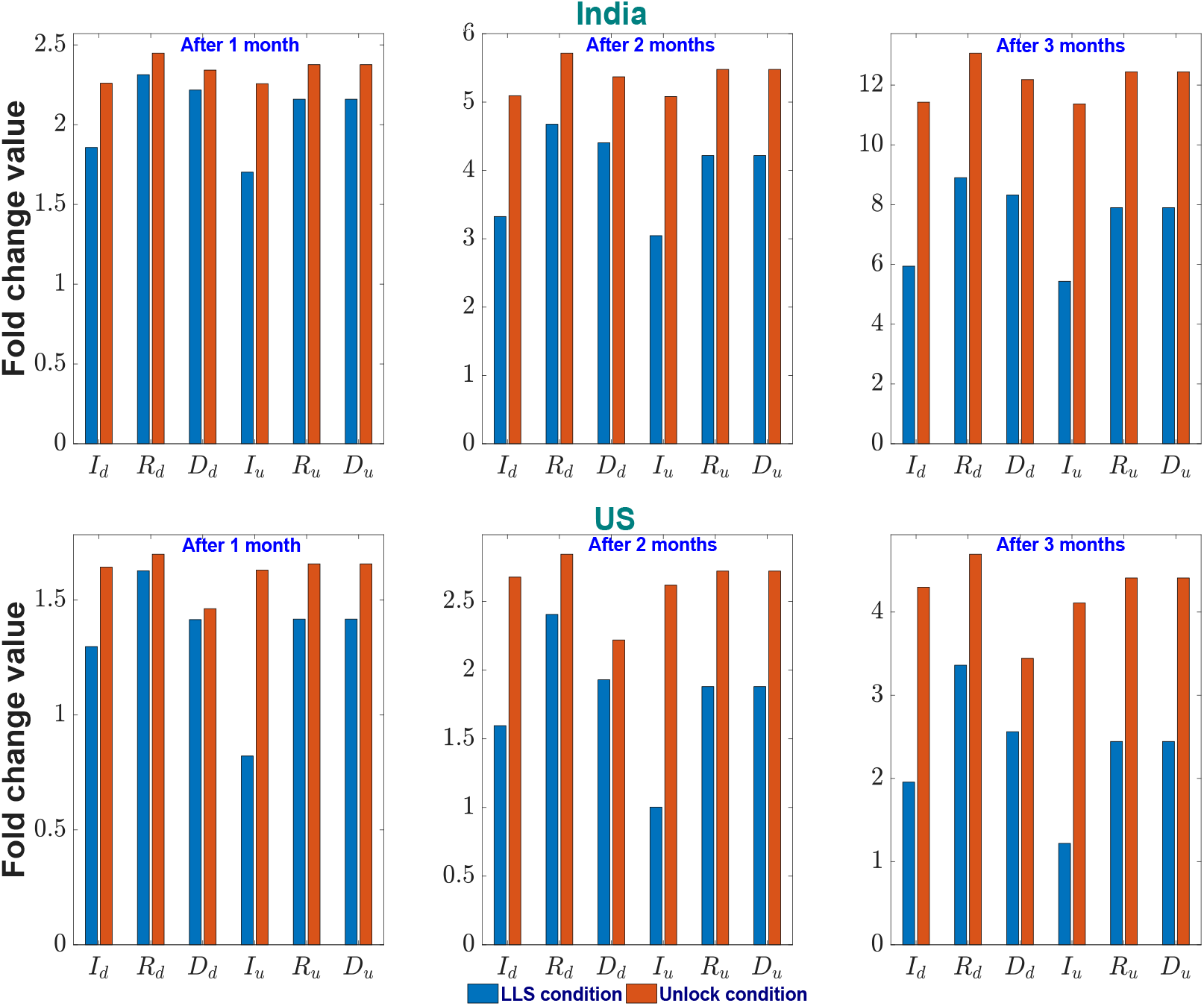
Fold change in the detected and undetected infectious compartments after one, two and three months under two different parametric situations. The red bars represent the fold change if parameters take the ULS values, and the blue bar represents the same if parameters take the LLS values (see Table 2). Fold change is measured taking July 27 value as their bases. The upper row represents the scenario for India and the lower row for the US.

## 5 Possible policy hypothesis through parameter recalibration

We look for policies that could be implemented during the unlock phase to restrict disease spreading. We used parameter recalibration techniques to define such policies that may be adopted to slow down the infection. To choose parameters for our recalibration exercise, global sensitivity analysis (Figure 8) was done to identify the parameters which have the potentiality to regulate the disease dynamics during the unlock period. It is to be noted that the detected class and the undetected class have different values for the parameters *β*_0_ and *ν* during the unlock period, simply because the former has no or limited movement and the later has free movement. We, therefore, considered different notations of *β*_0_ and *ν* for two groups. In particular, *β*_0 &_ *ν* were replaced by *β*_01 &_ *ν*_1_ for detected class, and the same for the undetected class by *β*_02 &_ *ν*_2_. Figure 8 shows that *α, β*_01_, *β*_02_, *ν*_1_ and *ν*_2_ all are sensitive parameters. However, these parameters cannot be considered for recalibration exercise because during the unlock period one does not have any control over 243 these parameters and the values for the undetected class are completely unknown. We, therefore, looked for other parameters which are sensitive and controllable. Figure 8 shows that the parameters *γ* and *ω*, having negative PRCC values, are also sensitive and they have more effect on the undetected class than the detected one. So, we selected *γ* and *ω* as the target parameters for our recalibration exercise to hypothesis on the controlling measures. It is mentionable that the parameters *γ* and *ω* are measurable and controllable. In fact, their values can be increased by increasing Covid-19 testing.

**Figure 8:**
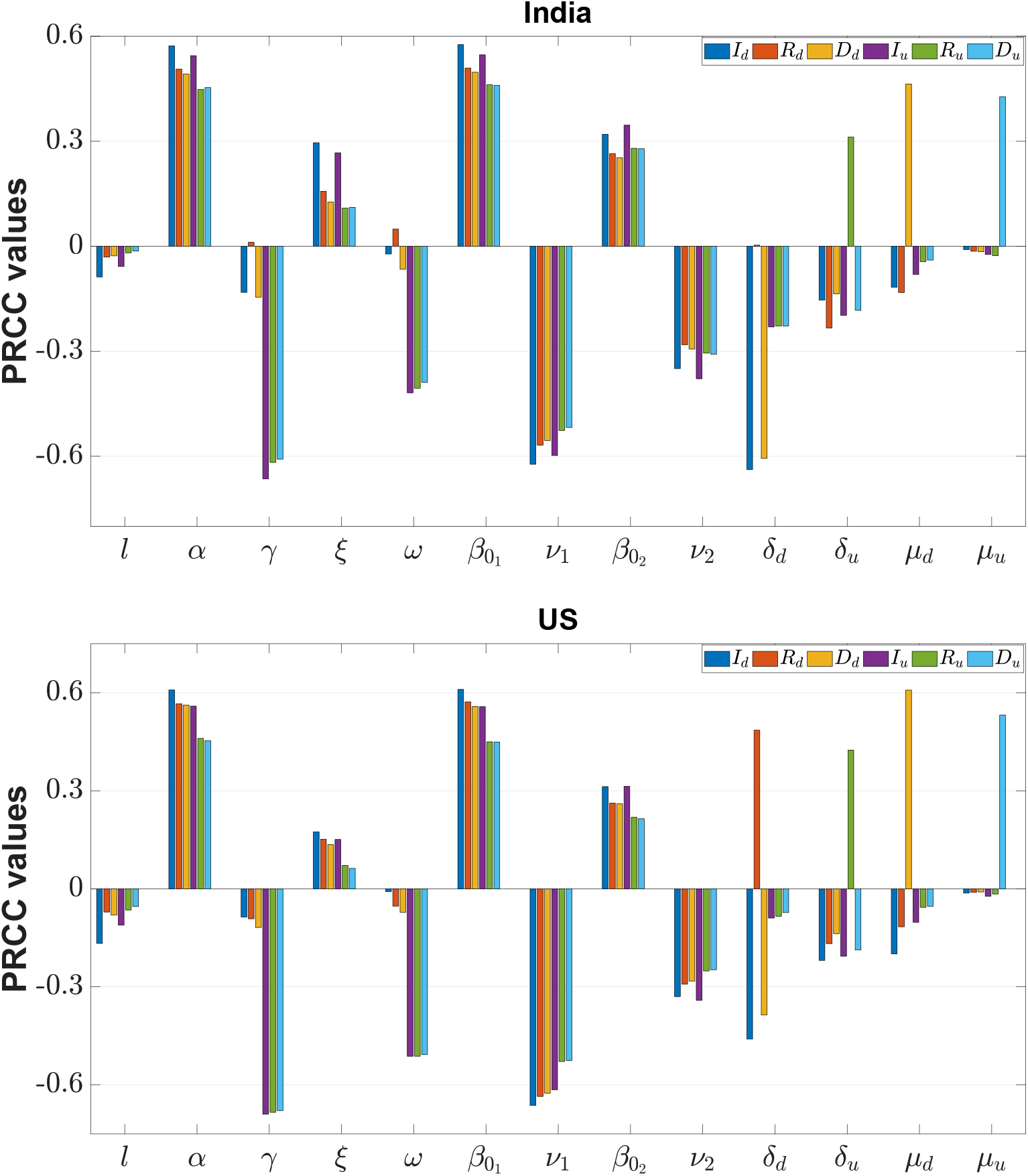
Sensitivity of Indian and US parameters during the unlock period. The parameters *β*_01_, *β*_02_ are the baseline disease transmission rates and *ν*_1_, *ν*_2_ are restriction-measuring parameters in the movement for detected and undetected infected classes.

After identifying the targeted parameters, we now observe how much these parameters can control the disease progression during unlock period. In this process, we increased separately the values of γ and ω by 25%, 50%, 75% and 100% from their existing values of unlocking 253 condition and estimated the fold change in the number of detected infected individuals (*I_d_*) and undetected infected individuals (*I_u_*) after 1, 2 and 3 months (see Figure 9). These fold change values were compared with the corresponding fold change values of *I_d_* and *I_u_* classes of Figure 7. Increased values of *γ* and *ω* reduced both the numbers of detected and undetected infected individuals significantly. For an example, if the value of *γ* is increased by 50% of the existing rate, India would be able to reduce the number of detected individuals by 10%, 30%, 40% and undetected infected individuals by 40%, 50%, 60% in the next one, two and three months, respectively and the value even can go below the corresponding value of LLS condition. We also noticed a higher variation in the undetected class, *I_u_*, compare to the detected class, *I_d_*.

**Figure 9:**
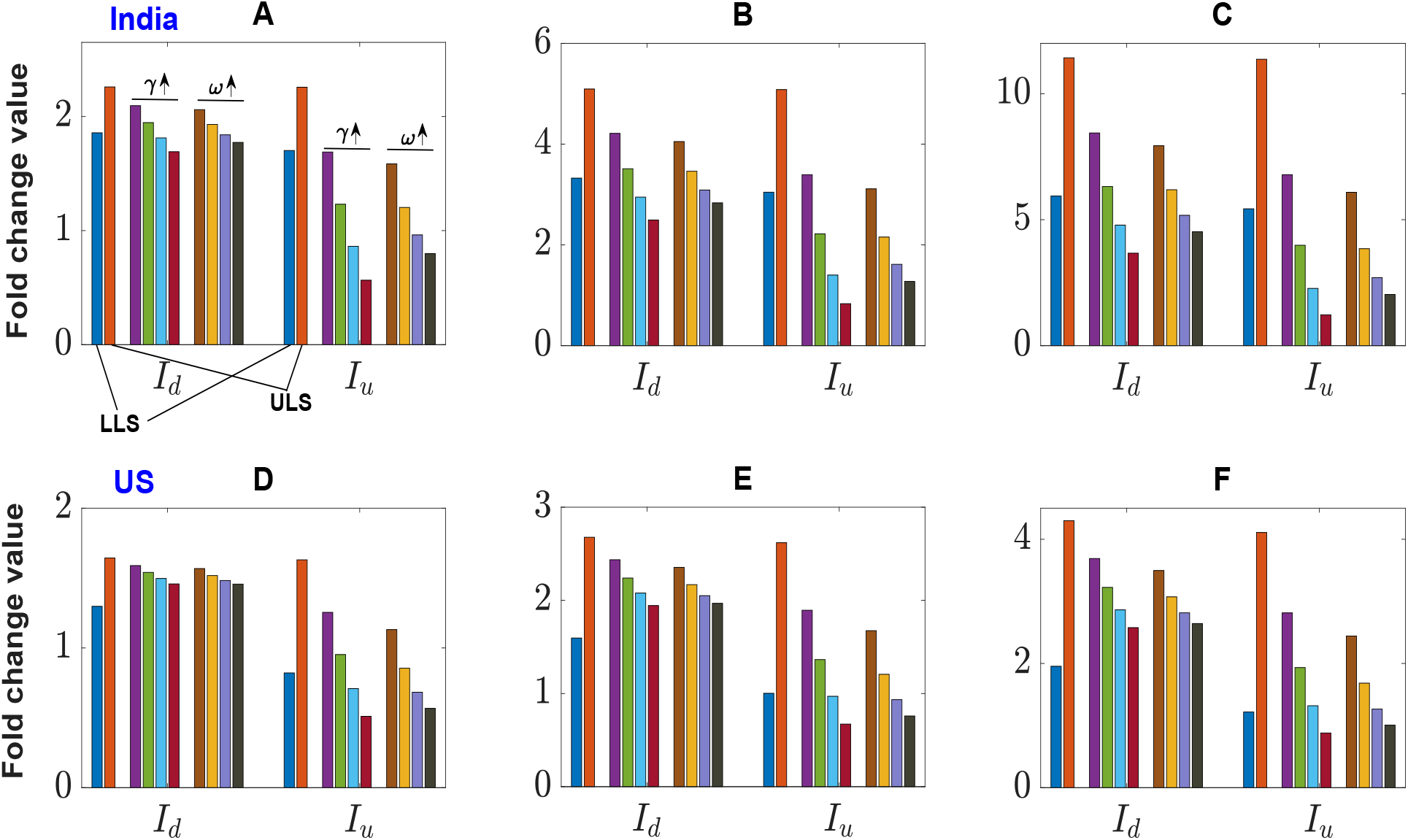
Parameter recalibration. Fold change in the detected infected individuals (*I_d_*) and undetected infected individual (*I_u_*) after one month (Fig. A), two month (Fig. B) and three months (Fig. C) due to an increment in the unlock values of *γ* and *ω* by 25%, 50%, 75% and 100%. Such fold changes have been represented by four consecutive bars under each parameter and compared with the corresponding fold change values of *I_d_* and *I_u_* classes of Figure 7, marked here by ULS and LLS. Fold change is measured taking July 27 value as their bases. The upper row represents the scenario for India and the lower row for the US.

Recent studies have shown that the use of repurposing drugs like dexamethasone, favipiravir and remdesivir can help the infected individuals to recover fast, and simultaneously decrease the disease-related death. The WHO has also recognized that the mortality rate of critically ill patients and can be reduced at least by one-third by using the repurposing drugs [52]. Countries like USA, UK and India have recently approved its use for the treatment of critically ill Covid-19 patients [53, 54, 55]. Here we investigated the effect of repurposing drugs on Covid-19 disease dynamics. Interestingly, we observed from our global sensitivity analysis that the death rate (*δ_d_*) and recovery rate (*µ_d_*) of the detected class are sensitive parameter, particularly for detected class (see Figure 8). 270 We, therefore, simultaneously increased the unlock value of *δ_d_* and decreased the value of *µ_d_* by 10%, 20%, 30%, 40%, and estimated the number of detected and undetected infected individuals after 1 month, 2 months and 3 months (see Figure 10). Simulation results clearly show a significant reduction in the number of detected infectives and undetected infectives. Also, in India, the number of detected and undetected infected individuals went below the corresponding values of LLS condition.

**Figure 10:**
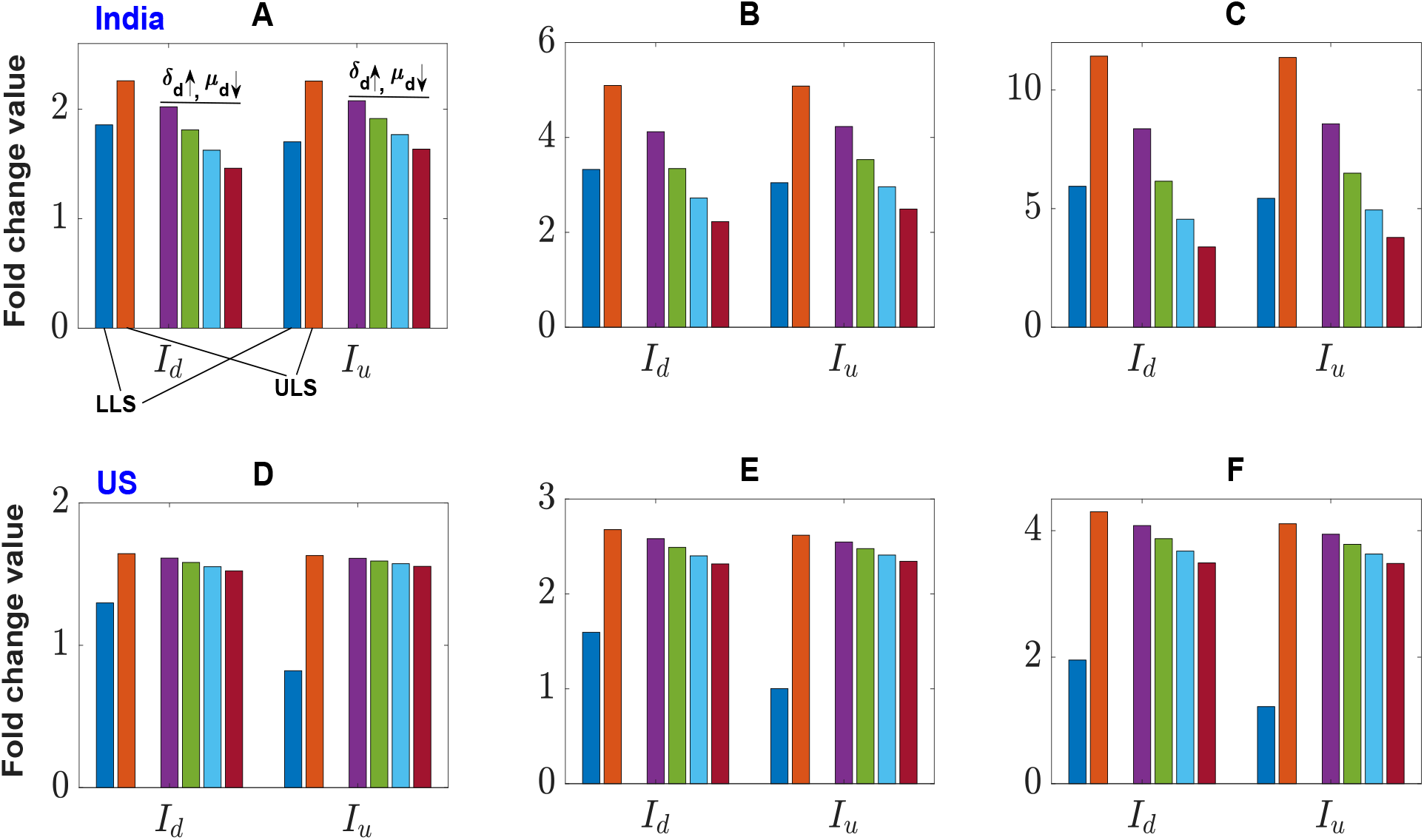
Effect of the repurposing drugs. Fold change in the detected and undetected infected classes after one month (Fig. A), two month (Fig. B) and three months (Fig. C) due to a simultaneous increment in the parameter *δ_d_* and a decrement in *µ_d_* in the unlock values by 10%, 20%, 30% and 40%. Such fold changes have been represented by four consecutive bars under each parameter and compared with the corresponding fold change values of *I_d_* and *I_u_* classes of Figure 7, marked here by ULS and LLS. Fold change is measured taking July 27 value as their bases. The upper row represents the scenario for India and the lower row for the US.

Finally, we observed the effect of perception strength (*l*) on the disease outcome. For this, we varied the parameter (*l*) from 0 to 100 and observed (Figure 11) the fold change in the detected and undetected infectives after 3 months with respect to its value as of July 27. This figure shows that there is a significant change in *I_d_* and *I_u_* classes in the case of India, however, the USA has a lesser effect.

**Figure 11:**
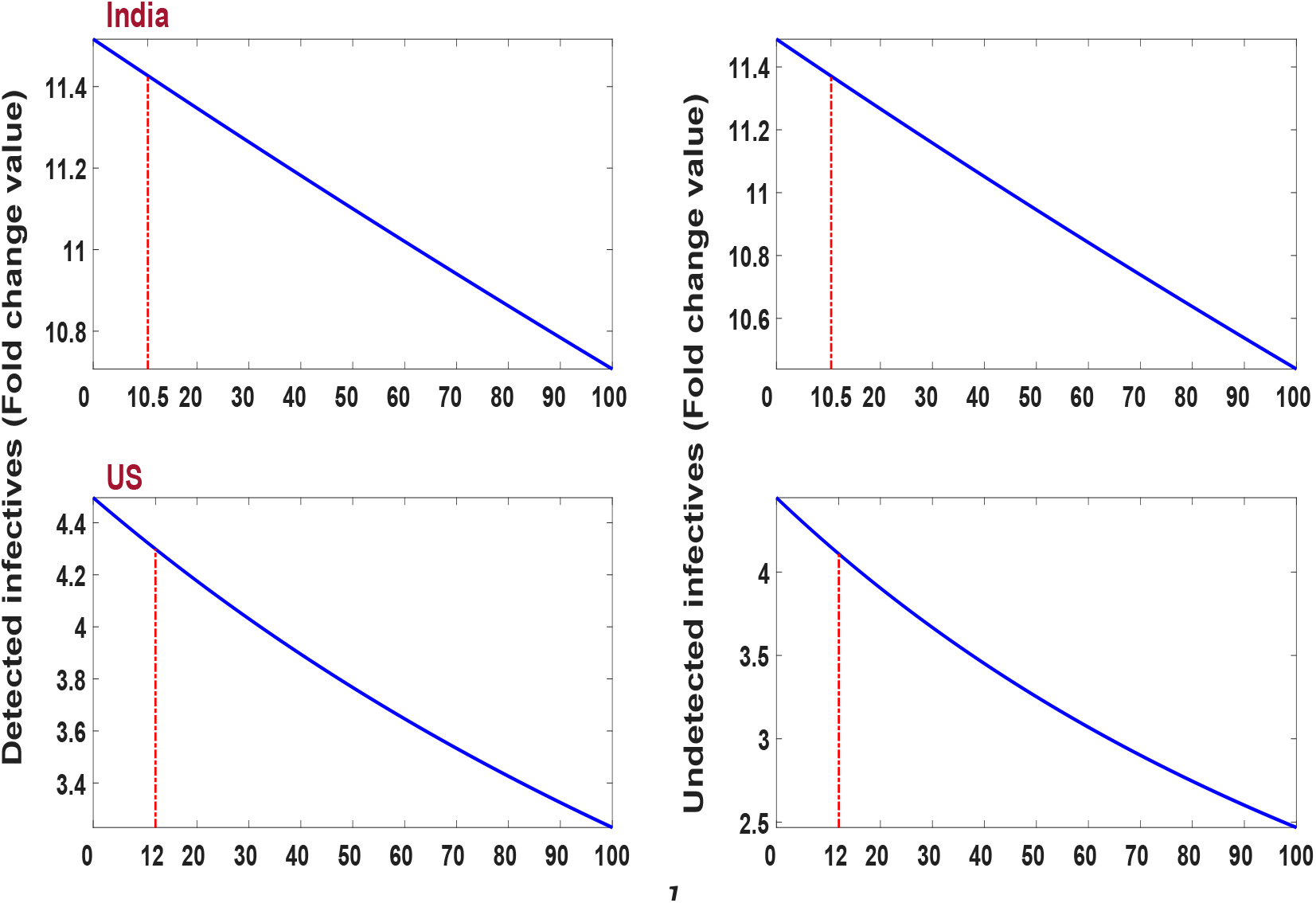
Variational effect of human perception on the levels of *I_d_* and *I_u_* classes after three months taking July 27 as their base values. It shows a significant change in *I_d_* and *I_u_* classes in the case of India when *l* varies in the range 0 to 100 (upper row). Such changes are little in the case of USA (lower row). The estimated value of *l* for India and USA are, respectively 10.5 and 12, which have been indicated by red lines. Other parameters are as in Table 2 (ULS).

## 6 Discussion

The world has now started to come out of the lockdown imposed due to Covid-19 pandemic even though the infection rate is still in an increasing mood in many countries. In the absence of any specific drug or vaccine, the main weapons to fight against the pandemic under unlock condition are remain the same, viz., maintaining social distancing & individual hygiene, compulsory use of mask, isolation of infectives or carriers. As the major portion of the infected individuals remains mildly symptomatic or asymptomatic, identification of such Covid-19 infected individuals is crucial for the containment of this deadly disease. This goal can be achieved if the potential individuals can be traced, tested and treated (3T principles) as quickly as possible so that the chances of spreading can be minimized. Though it may not be impossible, but definitely a hard task for many countries, where the population is large and/or medical facilities & resources are limited. Testing capacity around the world has become stretched due to a shortage of critical supplies, including test kits and face-masks, which may further decrease the covid testing and thereby increase the proportion of undetected cases. As the movement restriction is eased during the unlock period, this undetected class may enhance the disease spreading rate. So, the present study aims to assess the impact of undetected class on the Covid-19 dynamics during the unlock period. We began our study by understanding the effect of lockdown in the countries like Italy, Spain, 297 Germany and Switzerland, which have attained the epidemic peak and the curve is now going down. Gaining experience on the trend of data from these countries, we then studied the epidemiological status of countries like India and the USA, where the unlock phase has started even when the epidemic curve is going up. We proposed an extended SEIR epidemic model to address this problem. The disease transmission rate was modified to include (i) the effect of lockdown and (ii) public perception of getting an infection. It was presumed that the negative effect on the disease transmission due to human perception of getting the infection would increase with the disease-related death. Thus, the disease transmission rate, *β*(*t*), became a time-dependent parameter, which is usually assumed to be a constant in the epidemic models.

The data of the four countries, namely, Italy, Spain, Germany and Switzerland, were considered to estimate the parameter values and looked for the critical time point from where the effect of lockdown was observed. Global sensitivity analysis with an initial guess of the parameter set identified five parameters as the most sensitive. We further modulated these parameters to search for the critical time point for which our system shows a good fit of the actual data when compared between different lockdown stages. In this process, we partitioned the whole time frame into three parts: before the lockdown stage (BLS), early lockdown stage (ELS) and late lockdown stage (LLS) by two critical time-points. The lockdown phase was divided into two parts, e.g., ELS and LLS, to best fit the model with the observed data through optimization technique. The best fit parameter set such obtained showed little effect of lockdown on the parameter *β*, the disease transmission coefficient, and *ν*, that encapsulates the effect of lockdown, during the ELS, where the disease transmission efficiency and lockdown effect measuring parameters have values *β*_0_ *>* 0.2 and 0 *< ν <* 0.5. However, the lockdown effect was more prominent on these parameters during LLS, having values *β*_0_ < 0.16 and 0.5 *< ν <* 1. The basic reproduction number, *R*_0_, for each country was estimated to be less than unity with the LLS parameter set, indicating that containment measures were successful to reduce disease transmission. It is to be mentioned that this transmission potentiality measure (*R*_0_) is a community health indicator and used by the health care administrators to decide the scale of the containment measure needed against the infectious disease.

In countries like India and the US, the epidemic curve is still growing with little impact of lockdown. Our simulation results showed that there exists a critical time point for both the countries where the effect of lockdown has been observed, but it was insufficient to flatten the curve. Our model simulation captured the disease spreading scenario for these two countries for the period January 22 to July 27, 2020. We noticed that the disease transmission coefficient, *β*(*t*), and the lockdown effect measuring parameter, *ν*, both decreased in the LLS stage from to its previous stage. These decreased parameter values, however, reduced the transmission potentiality measure, *R*_0_, but could not pull it below 1. Its value remained above 1 for both the countries, indicating that the epidemic is still in its growing stage. Possible reasons for the curve to still grow despite the longer lockdown lie in the observation that the recovery rates for the detected class (*δ_d_*) were low for the US and the value of baseline disease transmission rate (*β*_0_) did not change much in India compare to other countries (see Table 2). Also to add, these two countries are gradually coming out of the lockdown phase by removing movement restriction and other social restrictions. Withdrawal of such restrictions would increase the dissemination of infection in the community level, where contact tracing would be impossible. We, therefore, attempted to restrict the disease transmission rate during the unlock period at the level of the LLE stage, which is much lower. To find out the strategy, we used the parameter recalibration technique, where the parameters are manipulated to see their effect on the model outcome. This exercise identified two controllable sensitive parameters *γ*, which represents the translation rate of exposed class to detected class, and *ω*, which represents the translation rate from undetected to detected infected class. Recall that γ is the fraction of the exposed class who join the detected class *I_d_* through the identification of SARS-COV-2 in their body and the remaining fraction (1 *− γ*) becomes the member of *I_u_* class without going through testing. The parameter *ω* represents the joining rate of detected class from the undetected class through testing at a later time. Thus, both parameters are measurable and their values are linearly dependent on the number of tests performed. So, we considered these two parameters for our recalibration exercise and observed that the increase in these two parameters could slow down the growth curve. These two parameters could be increased by increasing the tracing and testing of individuals who are at-risk, thereby increase the number of detected individuals. This would help to control the spreading of the disease as detected individuals have lower chance to infect others and would help to break the chain. Our simulation results indicated (see Figure 9) that India could reduce the cases by 25% at the end of three months if the value of these parameters is increased by 50%. This has been successfully tested in countries like South Korea and Hong Kong. These countries have proved that the control over coronavirus may be achieved without imposing lockdown by increasing the test facilities, reducing the time for tracing and giving treatment at the earliest.

We have also captured the significance of public perception of disease spreading and observed that it can significantly change the number of infectives. Such effects were observed in both the countries India and the USA despite their socio-economic differences. We have also observed the effect of repurposing drugs in reducing the fatality of the disease. As there is no vaccine and specific drugs for treating SARS-CoV-2 infected individuals, we have to fight against the virus with the available treatments and control strategies in the unlock period. Our study emphasizes that enhancing covid testing and using repurposing drugs, we can significantly reduce the epidemiological burden. We believe our proposed strategy obtained through a mathematical model will help to make a better policy for the unlock phase.

## Data Availability

All the data used in this study are publically available.

## Acknowledgements

The work is supported by SERB (Govt of India) under MATRICS Scheme, Ref. No. MSC/2020/000020.

